# Private Practice Infectious Disease: Metro Infectious Disease Consultants

**DOI:** 10.1101/2023.06.07.23290801

**Authors:** Russell Petrak, Robert Fliegelman, Nicholas Van Hise, Vishnu Chundi, Vishal Didwania, Alice Han, Brian Harting

**Affiliations:** Metro Infectious Disease Consultants, Burr Ridge, IL 60527, USA

## Abstract

Despite the stimulating nature of infectious diseases (ID), fellowship programs have failed, on a yearly basis, to fill multiple positions. In addition, numerous entities have published information showing that infectious disease physicians are poorly reimbursed compared to their colleagues in other specialties and likely to feel overwhelmed. To assess the status of our ID group and define opportunities for improvement, Metro Infectious Disease Consultants (MIDC) conducted an internal audit. Eighty-nine percent of employees and 94% of partners were satisfied or very satisfied with MIDC. In addition, based on a Medscape survey, MIDC employees received financial compensation that was 18% greater and partners 235% greater than the average for all ID physicians. A private practice model of infectious diseases is a viable, lucrative, and professionally stimulating option to other ID models. This allows for enhanced lifestyle flexibility and should be viewed by potential ID applicants as a realistic and widely available option to practice this dynamic specialty.

## Introduction

The practice of infectious disease (ID) is the most dynamic, stimulating, and challenging subspecialty in medicine. It interfaces with virtually every other medical and surgical specialty, allowing for a diverse patient experience. Hospitals rely on ID physicians to direct antibiotic stewardship and infection control programs, as well as therapeutic guideline development. Despite extensive efforts by the Infectious Disease Society of America (IDSA), these realities have not been imprinted on the medical workforce in training. Over the last 11 years (1), ID fellowships have failed to fill open positions. Surveys have consistently listed ID as one of the lowest paid specialties with one of the highest rates of burn out – a non-sustainable situation (1-4). The prosperity, lifestyle options, and overall satisfaction with the clinical practice of infectious disease has, unfortunately, been lost in this morass. In an attempt to identify unknown issues, Metro Infectious Disease Consultants (MIDC) conducted an internal physician audit.

## MIDC Overview

MIDC is comprised of 112 infectious disease physicians working in 26 offices, 110 hospitals and 8 states. Physicians are stratified by four levels: employee, partner, senior partner, and founding partner. Of the 106 ID physicians, 63 are male and 43 are female. Ages range from 31 to 70 with a median age of 46.

Multiple schedules are available for employees including full or part time, and a 7 days of consecutive work followed by 7 days off (7/7). In addition, MIDC employs 190 physician extenders. These professionals are largely utilized in the inpatient setting to clinically support a larger or more challenging patient population. This results in a more organized workflow, limiting the inherent inefficiencies of hospital care. Physician extenders are also invaluable in the outpatient arena for infusion therapy administration and management. In addition, the supervising physician has the opportunity to provide educational support for the extender.

One hundred and eighty-three nurses provide support for in-office patient care, OPAT evaluation, and infusion administration. Sixteen pharmacists are utilized for antimicrobial stewardship, clinical research, CME support, and medication safety.

Having achieved ACCME certification in 2017, MIDC awarded over 1500 hours of category 1 infectious disease CME to their physicians in 2022 alone. With the onset of the pandemic, MIDC formed a weekly conference available by teleconference. This expedited the transmission of rapidly changing information to the physician, extender, and pharmacy workforce. In addition, these educational events enhanced collegiality and solidified MIDC’s efforts to manage these critically ill patients.

MIDC revenue generation is diverse, including inpatient and outpatient consultations, outpatient parenteral antibiotic therapy (OPAT), biologic infusion supervision, clinical research, and contract support. MIDC also services numerous contracts for infection control, antibiotic stewardship, and medical leadership.

## Methods

An anonymous audit was sent by email to MIDC physicians. The founding partners, physicians who had joined the group less than 6 months before the audit, and any physicians whose status in the group has changed in the last 6 months were not included in the audit. Evaluation included degree of satisfaction stratified as very satisfied, satisfied, neutral, dissatisfied, and very dissatisfied.

Satisfaction was further analyzed by the region in which the physician worked, employee or partner, type of work schedule, average number of patients seen and average number of hospitals serviced per day. In addition, physicians were asked which aspects of the group resulted in the highest job fulfillment. Lastly, an internal financial evaluation was conducted by reviewing the total compensation for each physician and comparing this to the Medscape 2022 and the IDSA 2017 audits.

## Results

The audit was sent to 70 physicians and was completed by 67 (95.7%). Of the 67 respondents, 25 were employees, 42 were partners. Overall, 32 (48%) physicians were very satisfied, 28 (41%) were satisfied, 5 (8%) were neutral, and 2 (3%) were dissatisfied. 22 (52%) partners were very satisfied, 18 (42%) were satisfied, 1 (2%) was neutral, and 1 (2%) was dissatisfied. Ten (40%) employees were very satisfied, 10 (40%) were satisfied, 4 (16%) were neutral, and 1 (4%) was dissatisfied. Of the physicians working full time, 29 (52%) were very satisfied, 22 (39%) were satisfied, 3 (5%) were neutral, and 2 (4%) were dissatisfied. Two (33%) part-time employees were very satisfied, and 4 (67%) were satisfied. One (20%) of the employees working the 7/7 schedule was very satisfied, 2 (40%) were satisfied, and 2 (40%) were neutral.

The most valuable aspects resulting in job fulfillment varied slightly between partner and employed MIDC physicians. The 4 most valuable aspects for partners were financial stability 37 (88%), group collegiality 33 (79%), nursing/pharmacy support 27 (64%), and infrastructure support 24 (57%). Employees valued infrastructure support 22 (88%), financial stability 19 (76%), nursing/pharmacy support 16 (64%), and lifestyle flexibility 16 (64%).

Seventy-one (71%) percent of physicians serviced 1 hospital per day, while 21% serviced 2, 6% serviced 3, and 2% more than 3. Seventy seven percent (77%) of physicians saw less than 25 patients per day, with 36 (36%) seeing less than 20.

The average yearly remuneration for MIDC employees was 307 thousand dollars, while MIDC partners earned a yearly average of 610 thousand dollars.

## Analysis

MIDC physicians are highly compensated compared to other ID physicians based on other available audits (2-4). A Medscape survey in 2022 listed the yearly average for all ID physicians as 260 thousand dollars. The IDSA 2017 survey showed private practice ID physicians earning an average of 316 thousand dollars yearly. In contrast, the employed physicians of MIDC receive a total remuneration almost identical to the average of all private practice ID physicians. MIDC partners receive an average remuneration that is 93% higher than the average listed for private practice ID physicians in the same IDSA survey (2).

In addition, 90% of MIDC physicians audited are satisfied or very satisfied. While interpretation of this satisfaction may be complex, the listing of valuable MIDC aspects would suggest that financial stability is a key factor. ID conversations that deal with the value of infectious disease frequently discuss the ability to direct patient care, interface with multiple factions of a hospital and medical staff, and cost containment efforts. While these are noble and definitive skills, they are not well reimbursed or financially valued. MIDC optimizes reimbursement through a diversification of revenue which includes, but is not limited to, patient consultation. This provides a safety net as federal payers continue to implement payment cuts over the next several years. These factors in turn lead to MIDC’s salary numbers being dramatically higher for both employees and partners. The Medscape survey listed the average salary for an ID physician as 260 thousand dollars. In contrast, MIDC employees averaged 18% higher and partners 235% higher than those published numbers.

The ability to incorporate flexible work schedules, such as 7/7 and part time models, is attractive to a large segment of the ID work force – both men and women. Utilizing these models to accommodate work life balance has and should continue to assist in recruiting and retaining clinicians. MIDC employed physicians highly valued this flexibility, as it will help them accommodate to personal stresses while maintaining a professional career.

Clearly, MIDC values physician extenders and incorporates their skills widely. The utilization of extenders results in a clinician’s ability to service more patients while spending less time documenting the encounters. Physicians have the opportunity to continue to educate the extender and generate close working relationships resulting in enhanced efficiencies of care. Obviously, this translates to increased productivity and time for either professional development or personal endeavors.

As life-long learners, education is an anchor tenant for any ID practice, regardless of model, location, or size. In an environment where private practice ID physicians are not typically associated with progressive education, MIDC has invested significant time and money to refute this belief. MIDC utilizes ACCME certification to provide timely and pertinent infectious disease education to its physicians.

In short, MIDC is organized to optimize patient care by providing a strong clinical work force, surrounded by a robust infrastructure and financial stability. This, combined with lifestyle flexibility, allows for a diverse and collegial atmosphere, adaptable to the vast majority of infectious disease clinicians.

## Data Availability

All data produced in the present study are available upon reasonable request to the authors

